# *APOL1* variants affect APOL1 plasma protein concentration: a UK Biobank study

**DOI:** 10.1101/2024.02.28.24303461

**Authors:** Walt E. Adamson, Harry Noyes, John Ogunsola, Rulan S. Parekh, Anneli Cooper, Annette MacLeod

## Abstract

The *APOL1* gene is associated with chronic kidney disease progression. Circulating APOL1 protein levels are likely to play a critical role. Using UK Biobank data from 43,330 participants with APOL1 protein concentration levels, we found that individuals self-reporting as Black or Black British individuals had significantly higher serum APOL1 levels than all other ethnicities. To investigate the genetic factors underlying this difference, we explored the impact of APOL1 genotypes and other potential modifiers on circulating APOL1 protein levels. We analysed APOL1 protein concentration data in 1,050 UK Biobank participants of recent sub-Saharan African ancestry, focusing on the APOL1 G1, G2, and N264K variants. APOL1 concentration showed a clear genotype-dependent effect: individuals with the G0/G0 genotype had the lowest levels, heterozygotes (G0/G1 and G0/G2), had intermediate levels, and individuals with the G2/G2 genotype had the highest levels, demonstrating a dose-dependent relationship. The N264K variant reduced protein levels on a G2 background (p = 6 x 10^−5^). However, even after accounting for genotype, APOL1 protein levels in Black or Black British individuals was still higher than other ethnicities. A genome-wide association study on this population identified no genome-wide significant loci, other than the *APOL1* gene itself (p = 3 x 10^−155^) associated with APOL1 protein levels. Findings confirm APOL1 genotype as the major genetic determinant of circulating protein levels and provide new insights into the potential phenotypic effects of the G1, G2, and N264K variants.

## Introduction

Chronic kidney disease (CKD) disproportionately affects individuals of recent African ancestry (1). This excess risk is partly attributable to two independent variants of the apolipoprotein L1 (*APOL1*) gene: G1 and G2 (2), which are common in sub-Saharan Africa and its diaspora, but rare in other populations. G1 (amino acid substitutions S342G and I384M) and G2 (deletion of N388 and Y389) are found in the same domain at the C-terminus of *APOL1* and exhibit complete linkage disequilibrium: haplotypes with both G1 and G2 alleles are either very rare or absent. As a result, six combined genotypes with respect to *APOL1* G1 and G2 have been observed (Table 1).

**Table 1:** The six observed combined genotypes with respect to the G1 and G2 loci. Genotypes with G1 and G2 on the same haplotype are theoretically possible but have not been observed.

| Genotype Name | Haplotypes |  | Variant copy number | G1 copy number | G2 copy number |
| --- | --- | --- | --- | --- | --- |
|  | G1 locus | G2 locus |  |  |  |
| G0/<br>G0 | AT<br>AT | TTATAA<br>TTATAA | 0 | 0 | 0 |
| G0/<br>G1 | AT<br>GG | TTATAA<br>TTATAA | 1 | 1 | 0 |
| G0/<br>G2 | AT<br>AT | TTATAA<br>6 bp deletion | 1 | 0 | 1 |
| G1/<br>G1 | GG<br>GG | TTATAA<br>TTATAA | 2 | 2 | 0 |
| G1/<br>G2 | GG<br>AT | TTATAA<br>6 bp deletion | 2 | 1 | 1 |
| G2/<br>G2 | AT<br>AT | 6 bp deletion<br>6 bp deletion | 2 | 0 | 2 |

APOL1 also plays a crucial role in protecting humans from highly pathogenic trypanosome parasite, *Trypanosoma brucei* by forming pores in the parasite’s membrane, leading to osmotic imbalance and cell death. However, two subspecies of *Trypanosoma brucei, T. b. rhodesiense*and T. b. rhodesiense, have evolved mechanisms to resist APOL1’s action and so can cause the often-fatal disease, human African trypanosomiasis. The G1 and G2 variants of APOL1 are associated with protection from human African trypanosomiasis (3). In *T*.*b. gambiense* infections, G1 and G2 are associated with decreased and increased risk of severe disease, respectively, while carriage of the G2 variant protects against infection by *T*.*b. rhodesiense* (3). Despite the association of different genotypes with distinct phenotypes in human African trypanosomiasis, studies of the association between kidney diseases and *APOL1* have often grouped the G1 and G2 variants together as recessively ‘high-risk’ in any combination. Carriage of two such alleles (i.e. G1 homozygotes, G2 homozygotes, and G1/G2 compound heterozygotes) is associated with a spectrum of kidney and related conditions including focal segmental glomerulosclerosis, hypertension-associated kidney failure, and HIV-associated nephropathy(2,4–6).

Association studies examining APOL1 variants are often limited in statistical power and have not reported data consistently by haplotype. In a prior study using UK Biobank data, we previously highlighted differences in association between the two-variant *APOL1* genotypes with kidney phenotypes, and demonstrated that the compound heterozygous genotype, G1/G2 (carried by millions of people worldwide), is deleteriously associated with 26 different conditions spanning human health(7). In contrast G1/G1 and G2/G2 genotypes were only associated with kidney conditions. Our analysis exposed complexities in the relationship between *APOL1* risk alleles and disease that are not evident when two-variant *APOL1* genotypes (G1/G1, G1/G2, and G2/G2) are grouped together as a homogeneous risk category.

*APOL1* also contains additional variants that may modify its impact on the kidney and in response to trypanosome infections. One such variant, N264K, was initially identified in an individual with the G2/G2 genotype, who was infected with an atypical *T*.*b. gambiense* strain. Parallel *in vitro* experiments demonstrated a reduction in the ability of APOL1 to lyse trypanosomes if N264K was present (8). This reduced trypanolytic activity was mirrored by a reduction in cytotoxicity in kidney cells (HEK293) as a result of APOL1 expression when N264K was present alongside G1 and G2 (9). Subsequently, it was shown that N264K, when co-inherited with G2, also reduces the renal toxicity associated with the G1/G2 and G2/G2 genotypes(10,11).

Given the recessive inheritance and incomplete penetrance, it is not clear how APOL1 causes cell injury in CKD. It is likely that there are both genetic and environmental factors needed to drive disease. A minor fraction of APOL1 protein is expressed by the kidney and other tissues but the majority is produced by the liver and circulates bound in protein complexes as a component of plasma HDL. The contribution of circulating APOL1 to disease processes remains a point of investigation. Serum APOL1 levels were correlated with some pathologies such as sepsis and COVID-19 severity (12), whereas other studies found no correlation with kidney disease risk(13).

Here, we aim to identify drivers of APOL1 protein levels using Olink protein quantification data of samples from the UK Biobank (14,15). Previously a genome-wide association study screening for associations with protein concentration in the UK Biobank identified APOL1 G2 (alongside SNPs rs138477541 or rs1053865678 on chromosomes 3 and 5 respectively) as having an association with APOL1 concentration (15). In this study, we examine differences in circulating APOL1 protein levels across ethnic groups. We then examined the impact of *APOL1* variants on APOL1 protein concentration by first undertaking a scan for associations between APOL1 protein expression and SNPs in the 1 MB region around *APOL1* and then testing whether the resulting cis-pQTL were within microRNA binding sites that might regulate *APOL1* expression.

## Methods

### Study design and participants

The UK Biobank is a prospective cohort study of 502,460 adults aged 40 to 69 years at enrolment between 2006 and 2010 from 22 assessment centres across the United Kingdom(16). At the baseline study visit, participants underwent nurse-led interviews and completed detailed questionnaires about medical history, medication use, sociodemographic factors, and lifestyle in addition to a rage of physical assessments and provided blood and urine. The UK Biobank study was approved by the North-West Multi Centre Research Ethics Committee, and all participants provided written informed consent.

### UK Biobank self-reported ethnicity data

Self-reported ethnicity data was obtained from UK Biobank data field 21000. Descriptors of ethnicity used here (Asian or Asian British, Black or Black British, Chinese, Mixed, and White) represent the top-level descriptors of ethnicity used in data field 21000.

### UK Biobank Olink protein quantification data

Olink protein quantification was performed on 54,219 UK Biobank participants(15). *APOL1* protein quantification data is available for 43,330 of these. Olink data is expressed in the Normalized Protein eXpression (NPX) scale, enabling relative quantification of the same protein across multiple samples. As NPX values are calculated on a log2 scale, an NPX difference of 1 represents a doubling of protein concentration.

### Genotyping

*APOL1* genotypes were obtained from the UK Biobank, which used a custom Affymetrix array for the G1 (rs73885319) and N264K (rs73885316) alleles. G2 (rs71785313) genotypes were imputed by the UK Biobank as previously described(16). The study includes includes only participants with complete, unambiguous *APOL1* G1 and G2 genotype data.

### Statistical analysis

APOL1 protein concentration was tested for association with *APOL1* genotypes with, age, sex, and genetic principal components 1-4 as covariates. Linear regression as implemented in R was used to compare the effects of *APOL1* genotypes and individual risk alleles. All statistical tests were 2-sided, and p < 0.05 was considered statistically significant.

### Identification of regulatory SNPs associated with APOL1 NPX

The published study of SNPs associated with APOL1 NPX only reports a single SNP within each QTL (15). To identify additional SNPs that might regulate APOL1 NPX we tested for associations between 21,406 SNPs and APOL1 NPX. SNPs were from the UK Biobank Affymetrix genotype data and within 500 Kb upstream and downstream of *APOL1*. Association testing was performed using Plink2(17) with a general linear model(18), using age, sex, and the first four principal components of the relevant sample set as covariates. SNPs that were associated with APOL1 NPX (p < 5 x 10^-8^) were tested for linkage with each other. Linked SNPs were clustered into groups where each locus was linked to at least one other in the group using the NetworkX package in Python(19).

To understand the mechanism of APOL1 NPX regulation, we tested whether the SNPs that were associated with APOL1 NPX in the Black/Black British and White populations were also within microRNA binding sites. A list of microRNA binding sites was downloaded from mirDB(20), and each binding site was tested for the presence of SNP loci with significant associations with *APOL1* NPX using a custom local Perl script.

## Results

### APOL1 protein concentration varies by self-reported ethnicity

Among the 54,219 UK Biobank participants for whom Olink protein abundance data was available, the mean age at recruitment and blood sampling was 56.8 years, 54% were female, while 93.3% and 2.3% self-reported White and Black ethnicity respectively. Olink data is expressed in the Normalized Protein expression (NPX) scale, enabling relative quantification of the same protein across multiple samples. As NPX values are calculated on a log2 scale, an NPX difference of 1 represents a doubling of protein concentration. We compared APOL1 protein NPX across self-reported ethnicities as defined by the UK Biobank: Asian or Asian British (which includes Indian, Pakistani, and Bangladeshi ethnicities); Black or Black British; Chinese; Mixed; White. All comparisons between self-reported ethnicities yielded significant differences (<3x10^-5^), except for the comparison between Asian and Chinese ethnicities (p = 0.31) (Table 2, Figure 1). UK Biobank participants with self-reported Black or Black British ancestry had almost two-fold higher NPX of APOL1 than other ethnicities.

**Table 2:** APOL1 NPX values by ethnicity. Differences in NPX between populations were tested using a T-test. NPX values are calculated on a log2 scale.

| Self-reported ethnicity | Number in group | Mean APOL1 concentration (NPX) | p (vs Black/Black British) | p (vs Chinese) | p (vs Mixed) | p (vs White) |
| --- | --- | --- | --- | --- | --- | --- |
| Asian/Asian British | 969 | 0.15 | $<2 \times 10^{-16}$ | 0.31 | $<2 \times 10^{-16}$ | $<2 \times 10^{-16}$ |
| Black/Black British | 1,124 | 1.10 | - | $<2 \times 10^{-16}$ | $<2 \times 10^{-16}$ | $<2 \times 10^{-16}$ |
| Chinese | 147 | 0.11 | - | - | $5 \times 10^{-14}$ | $2 \times 10^{-5}$ |
| Mixed | 340 | 0.38 | - | - | - | $4 \times 10^{-10}$ |
| White | 48,965 | 0.09 | - | - | - | - |

**Figure 1:**
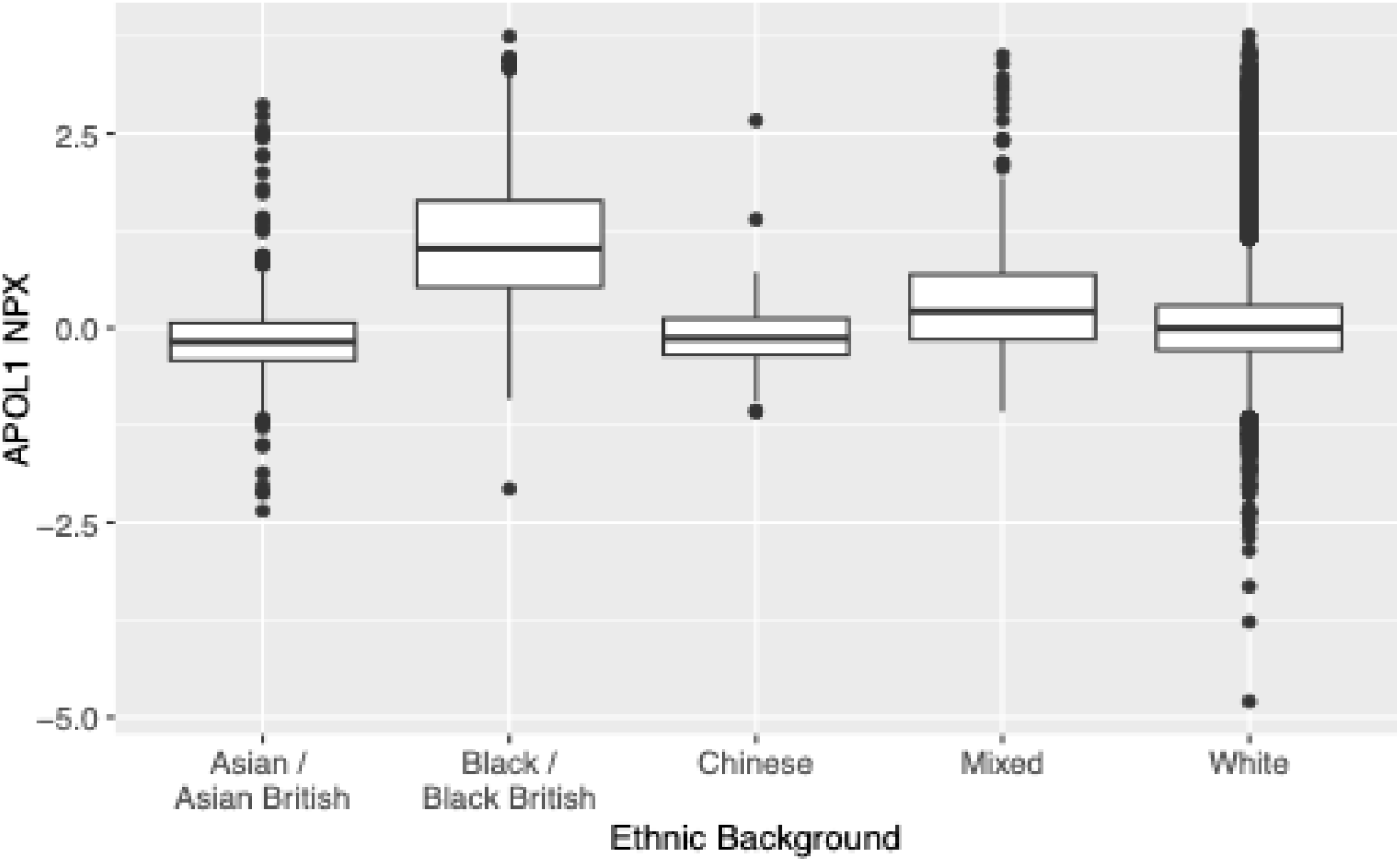
Distribution of APOL1 NPX for each self-reported ethnicity (as defined by the UK Biobank).

To begin exploring the genetic factors contributing to this observation, we investigated the impact of *APOL1* genotypes on circulating APOL1 protein NPX levels. Olink APOL1 protein NPX data and complete *APOL1* genotypes were available for 997 UK Biobank participants with self-reported Black or Black British ethnicity, among whom the mean age was 51.4 years and 54% were female.

### APOL1 protein NPX increases with number of G1 or G2 alleles

Previously, phenome-wide analysis highlighted differences in associations with health conditions between the six observed combined *APOL1* G1 and G2 genotypes(7), with two-variant APOL1 genotypes being associated with distinct kidney phenotypes, and the G1/G2 genotypes being uniquely associated with a range of conditions spanning human health. To investigate this further we examined the association of APOL1 protein NPX levels with each APOL1 genotype, revealing that among UK Biobank participants who self-report Black or Black British ethnicity, APOL1 NPX increases with the number of G1 and G2 alleles carried (Table 3, Figure 2). Compared to the wild-type (G0/G0) genotype, G2 homozygotes (G2/G2) had a 3.3-fold higher concentration. Linear regression confirmed the allele count effects: G1 alleles were associated with modest increases (OR 1.3, p = 2.8^-30^), while G2 alleles had stronger effects (OR 3.46, p = 7.6^-141^). Interestingly, individuals with the G0/G2 genotype, which is not associated with an increased risk of CKD, had higher APOL1 NPX levels than those with the G1/G1 genotype (t-test, p < 10^-16^), who are at increased CKD risk. Thus, disease risk associated with G1 and G2 alleles cannot be explained solely by plasma APOL1 concentration. The G1/G2 genotype appeared additive, with protein levels reflecting the combined effects of G1 and G2, suggesting that its broad phenotypic impact is not due to APOL1 concentration. Carriage of APOL1 G1 and G2 variants is extremely rare in populations that do not have recent sub-Saharan African ancestry, therefore it was not possible to assess the impact of G1 and G2 in other self-reported ethnicities.

**Table 3:** Associations between the number of *APOL1* G1 or G2 alleles carried and APOL1 plasma protein levels in the Black / Black British population.

| Grouping | Number in cohort | Mean APOL1 concentration (NPX) |
| --- | --- | --- |
| 0 G1 or G2 alleles (G0/G0) | 446 | 0.60 |
| 1 G1 allele (G0/G1) | 285 | 1.05 |
| 2 G1 alleles (G1/G1) | 78 | 1.40 |
| 1 G2 allele (G0/G2) | 136 | 2.11 |
| 2 G2 allele (G2/G2) | 21 | 2.89 |
| 1 G1 allele and 1 G2 allele (G1G2) | 31 | 2.37 |

**Figure 2:**
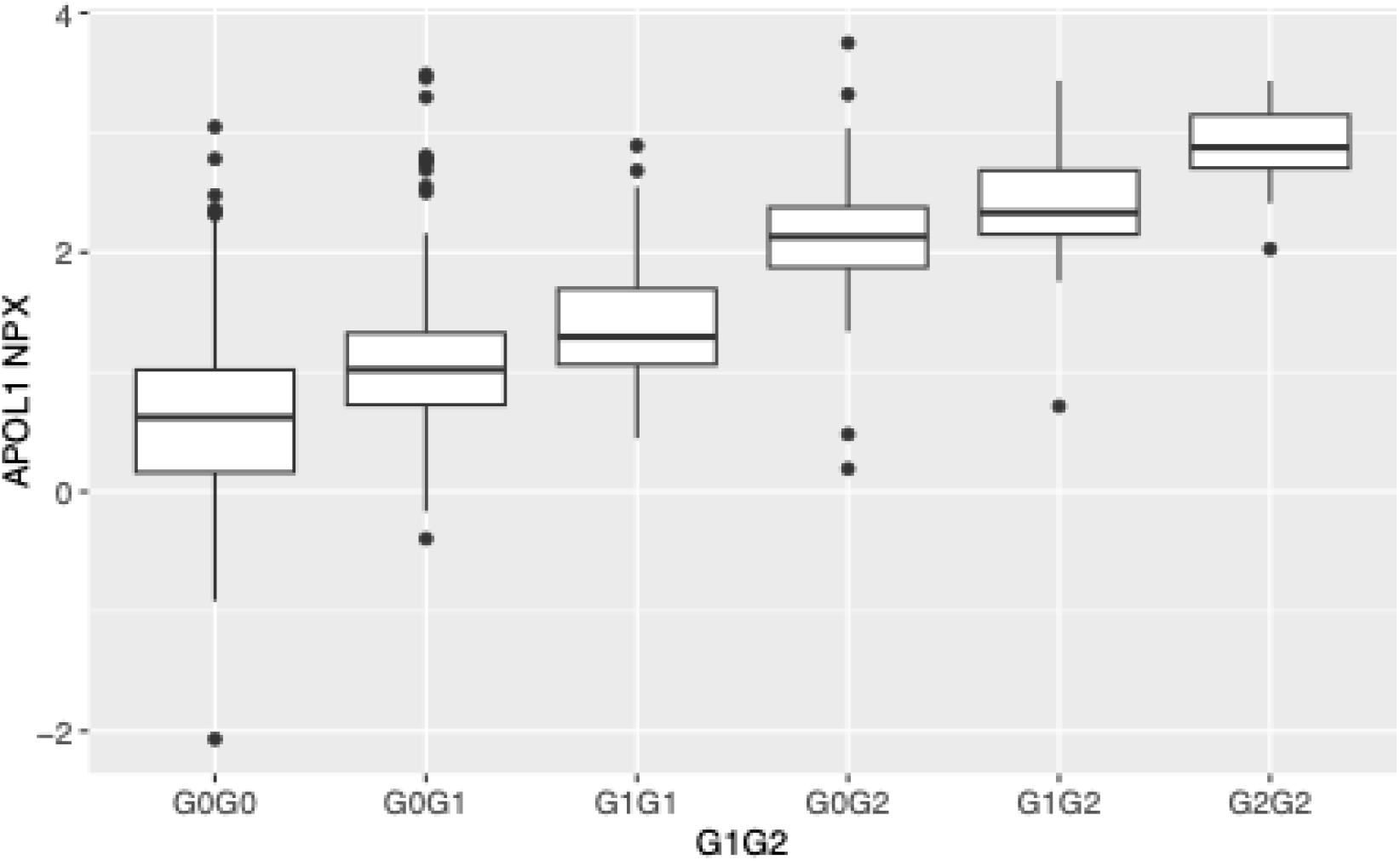
Boxplot showing distribution of APOL1 concentrations in the Black / Black British population with 0, 1, or 2 G1 or G2 alleles. Both G1 and G2 are associated with higher APOL1 NPX levels but G2 is associated with a much larger increase in APOL1 NPX.

### APOL1 protein NPX levels in the G0/G0 genotype varies by self-reported ethnicity

Among UK Biobank Black/Black British participants, mean NPX (1.10) is significantly higher (p < 2 x 10-^16^) than those who report other ethnicities (mean NPX 0.09) (Fig 1 Table 2). A proportion of this difference can be attributed to the overrepresentation of G1 and G2 alleles in this cohort, which are rare or absent in non-sub-Saharan African populations. However, among Black/Black British participants with the G0/G0 genotype, APOL1 concentration (0.61 NPX) was also 45% higher (p < 2x10^-16^) than the remainder of UK Biobank participants with that genotype (mean = 0.07 NPX), indicating that among populations with recent sub-Saharan African ancestry, additional loci and environmental factors might impact APOL1 concentration.

### Evidence that N264K is detected in G0 and G2 haplotypes, but not G1 haplotypes

UK Biobank participants with self-reported sub-Saharan African ancestry includes 6,862 individuals with an identifiable genotype at the N264 locus. It had previously been reported that the N264K variant occurs on G0 and G2 haplotypes as a result of two independent mutational events, and that N264K is mutually exclusive with the *APOL1* G1 allele(11). Our analysis of the UK Biobank data supports this observation: N264K was observed on a G0/G0 background in 77/2,639 individuals (2.9%), and on a G2/G2 background in 7/129 individuals (5.5%). N264K was not observed on a G1/G1 background 0/598 individuals (0.0%) (Table 4). Among individuals with the G0/G0 genotype, the presence of N264K had no effect on *APOL1* protein concentration (p = 0.88). Similarly, N264K had no effect on *APOL1* concentration on a G1 background (genotypes G0/G1, G1/G1, or G1/G2, p = 0.89) (Table 5).

**Table 4:** Distribution of N264K haplotypes across APOL1 genotypes.

| Genotype | Number in cohort | 0 N264K haplotypes | 1 N264K haplotype | 2 N264K haplotypes |
| --- | --- | --- | --- | --- |
| G0/G0 | 2,639 | 2,562 | 77 | 0 |
| G0/G1 | 2,069 | 2,026 | 43 | 0 |
| G0/G2 | 1,124 | 1,050 | 72 | 2 |
| G1/G1 | 598 | 598 | 0 | 0 |
| G1/G2 | 303 | 285 | 18 | 0 |
| G2/G2 | 129 | 122 | 7 | 0 |

**Table 5:**
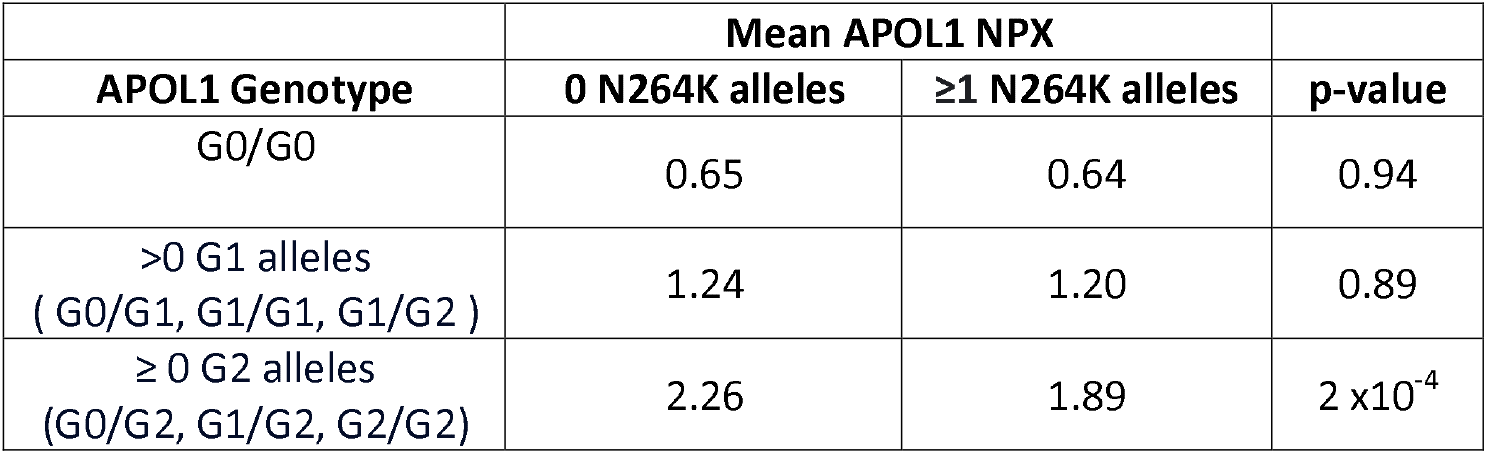
The effect of the N264K genotype on backgrounds containing: 0 G1 or G2 alleles; at least one G1 allele; at least one G2 allele. NPX values are calculated on a log2 scale.

### In a G2 background, N264K reduces APOL1 concentration

On a G2 background (G0/G2, G1/G2, or G2/G2), the presence of N264K was associated with significantly lower APOL1 protein level (t-test, p = 2 x 10^-4^) (Table 5). The mean APOL1 protein concentration was 17% lower among participants who carried N264K and at least one G2 allele than those who carried G2 alone.

### Loci that regulate APOL1 NPX in White and Black British populations

In the White British and Irish populations, eleven distinct protein Quantitive Trait Loci (pQTL) have been associated with APOL1 concentration (15) (Table S1). However, only three pQTL were identified in the Black/Black British population (Table S2). We examined these loci to determine whether they might contribute to the difference between sub-Saharan Africans and individuals of other ethnicities. At the two loci of largest effect in the White British/Irish population (Chr16, rs763665, OR 1.7; Chr22, rs2239785, OR 1.4), the allele associated with higher NPX was more common in Black/Black British than White British/Irish participants (Table S1). This could contribute to the higher APOL1 NPX in the Black/Black British group. The effects of the remaining nine loci were very small: some increased and some reduced APOL1 NPX in the Black/Black British population relative to the White British/Irish population (Table S1) and the combined effect of these nine loci on population level APOL1 NPX is likely to be negligible. Interferon gamma (IFN-γ) has been shown to stimulate APOL1 expression(21), but in a linear regression model with sex and age covariates there was no evidence for IFN-γ NPX being associated with APOL1 NPX in the Olink data (p = 0.7).

### microRNA binding sites are associated with APOL1 concentration in the Black British population

The published list of SNPs associated with Olink APOL1 NPX only includes two in the *APOL1* region; rs2239785 and rs1317778148 in the White British/Irish and Black/Black British populations respectively(15). In order to identify SNPs that might regulate APOL1 NPX we undertook an association study of all SNP within 500 Mb of the start and end of *APOL1*. 154 SNPs in 13 linkage groups (r^2^ > 0.5) were associated with APOL1 NPX in the White British/Irish population with p < 5x10^-8^ (Table S3) and 95 in 10 linkage groups in the Black/Black British population (Table S4). The large number of linkage groups suggest that multiple loci are independently regulating APOL1 NPX. In the White British/Irish population, the mean odds ratio was only just over 1 (mean 1.03, 95% CI 1.015-1.044), whilst in the Black British population it was 1.49 (95% CI 1.38-1.59) suggesting much stronger cis regulation of APOL1 NPX in the latter population consistent with the much higher *APOL1* NPX in this group.

A list of microRNA binding sites in the 3’ untranslated regions of APOL1 was downloaded from mirDB(20), and each binding site was tested for the presence of SNP loci with significant associations with APOL1 NPX (p < 5 x. 10^-8^). Associations with each SNP in each predicted microRNA binding site are shown in Table S5, and a summary is shown in Table 6. In the Black/Black British population, two linked SNPs (22:36663535_CTG_C and 22:36662950_CA_C) overlapped with 4 and 7 miRNA binding sites respectively and were strongly associated with higher APOL1 NPX (OR 1.95 and 1.89 respectively). Both SNPs are in linkage disequilibrium with G2 (r^2^ = 0.58 and 0.51 respectively), suggesting altered miRNA binding may contribute to the elevated APOL1 expression associated with this allele. Remarkably, in the White British population, the same two SNPs were associated with small reductions in APOL1 NPX despite similar minor allele frequencies (Table 6). It is possible that these SNP interact with different miRNA in the two populations. An additional nine SNPs in two linkage groups in the 3’ UTR were associated with lower APOL1 levels in the white British population. Collectively, these findings suggest that variation in miRNA binding sites contributes to population differences in APOL1 NPX and may provide a mechanism linking G2 to higher protein abundance.

**Table 6:** SNP associated with APOL1 NPX and in microRNA binding sites of APOL1 3’UTR in the Black/Black British and White populations. Count miRNA indicates the number of different miRNA that have binding sites that cover that SNP. Odds ratio and p-values for the association between the SNP and APOL1 plasma concentration are shown. Freq represents the frequency of the minor allele. See Table S3 for additional details of miRNAs at each SNP position.

|  |  | White British |  |  |  | Black British |  |  |  |
| --- | --- | --- | --- | --- | --- | --- | --- | --- | --- |
| rs id | Position in 3' UTR | Count miRNA | OR | Freq | P | Count miRNA | OR | Freq | P |
| rs9610472 | 292 | 3 | 0.94 | 0.14 | $1.4^{-20}$ | 0 | NA | NA | |
| rs9610473 | 298 | 3 | 0.94 | 0.14 | $5.8^{-21}$ | 0 | NA | NA | |
| rs9610474 | 307 | 11 | 0.95 | 0.14 | $2.0^{-20}$ | 0 | NA | NA | |
| rs9610475 | 598 | 1 | 0.95 | 0.17 | $1.0^{-20}$ | 0 | NA | NA | |
| rs9610476 | 600 | 1 | 0.95 | 0.17 | $1.5^{-20}$ | 0 | NA | NA | |
| rs5750246 | 669 | 4 | 1.12 | 0.24 | $7.5^{-112}$ | 0 | NA | NA | |
| rs150349831 | 745 | 5 | 0.91 | 0.02 | $2.4^{-10}$ | 0 | NA | NA | |
| 22:36662950_CA_C | 871 | 7 | 0.96 | 0.22 | $5.2^{-15}$ | 7 | 1.89 | 0.24 | $9.4^{-52}$ |
| rs78523 | 1169 | 1 | 1.17 | 0.17 | $2.6^{-170}$ | 0 | NA | NA | |
| rs1142542 | 1254 | 4 | 0.95 | 0.17 | $4.1^{-24}$ | 0 | NA | NA | |
| 22:36663535_CTG_C | 1456 | 4 | 0.95 | 0.18 | $6.9^{-21}$ | 4 | 1.95 | 0.20 | $1.9^{-64}$ |

## Discussion

Recent laboratory and association studies have transformed our understanding of the human APOL1 gene: widening the scope of potentially detrimental effects in current populations as an evolutionary cost of surviving historic sleeping sickness epidemics (7,12,22–24) as well as identification of graded and differential risk by alleles, number of risk alleles, and also the presence of a modifying variant that mitigates APOL1 impact both on trypanosomiasis and CKD(8–11). Here, we further extend those findings to demonstrate that APOL1 G1 and G2 are both associated with dose dependent increases in APOL1 serum protein concentration, however the effect of G2 is far larger (Fig 2).

In studies examining the role of APOL1 in trypanosomiasis, the different effects of the two variants are well-established: G1 is associated with protection against severe disease in *T*.*b. gambiense*; G2 is associated with protection from *T*.*b. rhodesiense* infection, but is associated with increased disease severity in *T*.*b. gambiense* (3). Accordingly, trypanosomiasis studies routinely consider G1 and G2 separately. In contrast, CKD studies have typically grouped G1/G1, G1/G2, and G2/G2 together as ‘high-risk’. Recently, we demonstrated that this approach masks a deeper level of complexity, and identified conditions and phenotypes within CKD and beyond that were associated with particular two-variant *APOL1* genotypes(7). Here we extend these findings by demonstrating genotype-specific associations with APOL1 levels, and their modifiers.

The role APOL1 G2 in trypanosome infections is well documented. In *T*.*b. rhodesiense*, the parasite serum resistance-associated (SRA) protein binds APOL1 and blocks APOL1-mediated trypanolysis. However the two amino acid deletion present in the G2 variant modifies APOL1 to prevent SRA binding, restoring APOL1’s lytic function and conferring protection (25). Similarly, G2-specific phenotypes have been observed in CKD, with the G2/G2 genotype being associated with reduced (< 60 mL/min/1·73m^2^) estimated glomerular filtration rate(7). The mechanisms resulting in APOL1-mediated cell injury in CKD are unclear: multiple pathways have been proposed, and studies have typically examined genotypes G1/G1, G1/G2, and G2/G2 collectively as ‘high-risk’. If the causal mechanisms of CKD differ between G1 and G2, treating the variants as equivalent in studies will hinder efforts to further understand the pathogenesis of CKD.

Phenome-wide analyses also reinforce this point. We previously found the G1/G2 genotype to be associated with multiple deleterious conditions (as defined by *International Classification of Disease* (ICD) codes) (7), while detecting no associated with G2/G2 (either protective or deleterious). This was unexpected given that in mouse BAC transgenic models with the human promoter regions, G2/G2 has been shown to be associated with more severe kidney phenotypes and that this effect is dose dependent(26). The absence of associations with G2/G2 in our phenome scan could reflect reduced statistical power given the relative rarity of G2/G2. However the effect size for G2/G2 was smaller than that of the index genotype with a significant association in all cases and the odds ratio for the G2/G2 association was within the 95% confidence interval of the odds ratio for the index association in only 8 out of 27 associations of our previous phenome wide screen (Table S6) (7). This indicates that although lack of power might partially explain the lack of associations with G2/G2, lack of power is likely to be minor reason for the absence of associations. Therefore, either the G2/G2 genotype is not a risk factor for many conditions mediated by mechanisms other than CKD, or the effects of G2 homozygosity are masked by such factors as tissue specific concentration, altered substrate binding, protein folding variation or other biochemical properties that drive disease risk.

The N264K variant further illustrates the complexity of APOL1 biology. In a G2 background, N264K reduces trypanolytic function (8) and appears to protect against G2-associated kidney disease by disrupting APOL1 G2 pore-forming and ion channel conduction(10,11). Here, we show that in the presence of G2, the N264K variant also reduces APOL1 protein concentration. Although genotypes containing G2 and N264K have higher APOL1 protein concentration than G0/G0, the combined effects of reduced protein concentration and pore disruption might reduce the severity of the phenotype. N264K may also have additional, as yet unidentified, phenotype-modifying effects. The presence of both N264K and G2 in an individual has potential clinical implications: the G2 carriers without N264K are potentially at higher risk of developing kidney disease and could benefit from closer monitoring.

The data presented here also highlights additional contributors to APOL1 expression. Even among G0/G0 individuals, those with sub-Saharan African ancestry had significantly higher APOL1 NPX than individuals with other ethnicities (Figure 1). This indicates that APOL1 protein concentration is influenced by additional factors. MicroRNAs suppress expression of their target proteins and we have shown that microRNAs may contribute to the difference in APOL1 abundance between white and black British populations. The difference might also be attributable to many loci of small effect as well as environmental factors that were not incorporated into our model. The identification of additional loci and environmental factors that influence APOL1 protein concentration, and an assessment of their impact on human health would further develop our understanding of the protein and inform diagnostics and management for APOL1-related conditions.

There are some limitations to the study. The analysis is cross-sectional, and we cannot determine the longitudinal independent association with CKD and circulating APOL1 protein levels do not necessarily reflect tissue levels of APOL1 that could be more important for kidney damage. Our main strength is the large, well categorised study population, which enabled us to quantify population- and genotype-specific differences in APOL1 protein levels with precision.

## Conclusions

This study demonstrates the relationship between *APOL1* variants and circulating APOL1 protein levels. It identifies further phenotypic differences between the G1 and G2 variants, highlighting the value in considering them as distinct in molecular and association studies. Furthermore, it provides further detail on the relationship between the G2 and N264K variants, which has significant implications for diagnosis and therapy in kidney disease.

## Data Availability

This research has been conducted using data from the UK Biobank, a major biomedical database.

http://www.ukbiobank.ac.uk

## Data sharing statement

This research has been conducted using data from the UK Biobank, a major biomedical database: www.ukbiobank.ac.uk.

## Acknowledgements

We thank NHS England (Copyright 2023, NHS England). Re-used with the permission of the NHS England. All rights reserved. This work uses data provided by patients and collected by the NHS as part of their care and support.

We thank Public Health Scotland. (This research used data assets made available by National Safe Haven as part of the Data and Connectivity National Core Study, led by Health Data Research UK in partnership with the Office for National Statistics and funded by UK Research and Innovation).

We acknowledge the support of Dr. Richard Gregory and the Centre for Genomic Research (CGR) at the University of Liverpool for providing access to computational resources used in this research.

This study was funded by the Wellcome Trust (209511/Z/17/Z), H3Africa (H3A/18/004), and the Medical Research Council (UKRI434).

